# Intensive Community Care Services for Adolescents with Acute Psychiatric Emergencies: Trial Feasibility Findings from the Pilot Phase of a Multi-centre Randomised Controlled Trial

**DOI:** 10.1101/2025.07.12.25331424

**Authors:** Dennis Ougrin, Thilipan Thaventhiran, Ben Hoi-Ching Wong, Izabela Pilecka, Sabine Landau, Sarah Byford, Petrina Chu, Hassan Jafari, Margaret Heslin, Emma Tassie, Paula Reavey, Toby Zundel, Mandy Wait, Ruth Woolhouse, Tauseef Mehdi, Jovanka Tolmac, Joe Clacey, Leon Wehncke, Veronika Beatrice Dobler, Rhys Bevan-Jones

**Author notes:** Correspondence: Thilipan Thaventhiran. Wolfson Institute of Preventive Medicine. Queen Mary University London, Charterhouse Square, London EC1M 6BQ.

## Abstract

**Background:** Adolescents experiencing psychiatric emergencies often require intensive intervention to avoid hospital admission and support their transition into education, employment, or training (EET). Intensive Community Care Services (ICCS) offer a potential alternative to inpatient care. This pilot study aimed to assess the feasibility of conducting a randomised controlled trial (RCT) to evaluate the effectiveness of ICCS compared to treatment as usual (TAU) in reducing time to start or return to EET.

**Methods:** A multi-centre, parallel-group, single-blinded randomised controlled trial (RCT) with an internal pilot phase was conducted across seven NHS Trusts in the UK. Adolescents aged 12–17 experiencing psychiatric emergencies were randomised to ICCS or treatment as usual (TAU). The primary outcome was time to start or return to EET within six months. Secondary outcomes included clinical symptoms, functioning, service satisfaction, service use, and health-related quality of life. Descriptive statistics and hazard ratios were calculated to explore group differences. Feasibility was assessed against pre-specified progression criteria.

**Results:** Thirty-six adolescents were randomised during the internal pilot phase. The recruitment rate did not meet the progression criteria, and continuation to a full evaluation trial was deemed unfeasible. During follow-up, 83.3% (n=30) returned to EET, with a median time to EET of nine days (IQR: 1–49). Median time to EET was shorter in the ICCS group (6 days) compared to TAU (12 days), with a hazard ratio of 1.34 (95% CI: 0.63–2.86). ICCS was associated with improved service satisfaction, clinical symptoms, and functioning. The average cost per participant was higher in the TAU group (£15,155, SD 31,560) compared to ICCS (£7,063, SD 10,605), with minimal differences in quality-adjusted life years (QALYs). Fourteen safety events were reported across both groups.

**Conclusions:** A full evaluation trial was not feasible due to recruitment challenges, primarily arising from limited-service capacity to deliver two treatment pathways concurrently. Despite this, ICCS showed promising trends in clinical and functional outcomes and may offer a viable alternative to inpatient care. Further research is needed to explore the implementation and effectiveness of ICCS in routine practice.

**Trial registration:** ISRCTN, ISRCTN42999542. Registered 29/04/2020, https://doi.org/10.1186/ISRCTN42999542

## Background

Mental health crises among children and young people (CYP) are a significant public health concern. A recent National Health Service (NHS) survey in England revealed that approximately 1 in 6 children aged 5–16 years are likely to have a mental health disorder (1). In 2021-2022, over 1.2 million CYP were referred for mental health support, marking a 41% increase from the previous year (2). While Tier 4 Child and Adolescent Mental Health Services (CAMHS) inpatient units play a critical role in stabilising severe psychiatric conditions, they often result in prolonged admissions, with an average hospital stay of 120 days across all psychiatric units (3). Repeated and lengthy admissions can lead to interpersonal disconnection and increased strain on the healthcare system, exacerbating the challenges of providing timely, effective mental health care for young people (4).

Intensive Community Care Services (ICCS) for children and adolescents with severe psychiatric disorders, including home treatment, crisis teams, day services, and other intensive treatment teams, have been prioritised by national policy and commissioners in many countries (5). According to the NHS Long Term Plan, all NHS trusts must provide a form of ICCS in England by the end of 2024 (6). Despite this mandate, there is minimal evidence for the efficacy or effectiveness of ICCS (7). Recent systematic reviews indicate that studies of ICCS are highly heterogeneous, with varying outcome measures and inconsistent comparisons (8). Most studies have compared ICCS with inpatient care rather than other community-based services, limiting the understanding of its true effectiveness in real-world settings (9). Given these gaps in the evidence base, there is an urgent need for robust research on the clinical and cost-effectiveness of ICCS. The Comparison of Effectiveness and Cost-effectiveness of Intensive Community Care Services versus Treatment as Usual Including Inpatient Care for Young People with Psychiatric Emergencies (IVY) trial aimed to evaluate the impact of ICCS on time to return to or start education, employment, or training (EET), a key indicator of long-term recovery for CYP (10), as well as on a range of secondary outcomes, including psychopathology, functioning, and service satisfaction (11). The study design included an internal pilot phase to assess the feasibility of conducting a full evaluation trial. The study was to progress to a full-scale trial if pre-defined recruitment targets were met (12). In this paper, we report on the findings of the internal pilot.

## Methods

### Study Design

This study is a multi-centre, parallel group, randomised controlled trial (RCT) designed to evaluate the clinical effectiveness and cost-effectiveness of ICCS for young people experiencing psychiatric emergencies. The trial incorporated an internal pilot phase to assess the feasibility of recruitment before determining whether to proceed to a full-scale trial. The study compares ICCS with treatment as usual (TAU) across NHS Trusts in England. The trial followed CONSORT guidelines and received ethics approval. This pilot study is reported in accordance with the CONSORT 2010 extension for pilot and feasibility trials—a reporting framework rather than a methodological design guide—to ensure transparent presentation of feasibility objectives, process outcomes, and pilot-specific considerations.

### Participant Selection

Inclusion criteria required that participants be young people aged 12 to 17 years assessed by a consultant psychiatrist—typically within CAMHS crisis teams, paediatric wards, or emergency departments—as meeting clinical criteria for inpatient psychiatric admission or ICCS in participating NHS Trusts. To qualify for inpatient admission, at least one of the following had to be present: (1) acute suicidality requiring 24-hour observation; (2) recent medically significant suicide attempt; (3) new-onset or exacerbated psychosis; (4) severe affective dysregulation unmanageable in the community. Clinical eligibility for ICCS was determined by a consultant psychiatrist or senior ICCS clinician using all of the following criteria: (1) clinical stability suitable for intensive community care (i.e., absence of active psychosis or severe affective dysregulation requiring inpatient supervision); (2) CGAS score ≥ 20; (3) no acute risk necessitating 24-hour observation. Participants needed to demonstrate the ability to provide informed consent (or assent with parental consent for participants under 16 years). Exclusion criteria included individuals unable to consent due to mental state, those at a risk level incompatible with community care (CGAS score <20), and participants who could not be enrolled because local ICCS or TAU teams had reached full capacity (12). Diagnoses of Borderline Personality Disorder and Autism Spectrum Disorder were extracted from participants’ electronic clinical records, based on routine assessments by senior CAMHS clinicians; no additional trial-specific diagnostic instruments were used.

### Randomisation and Blinding

Participants were randomised in a 1:1 ratio to ICCS or TAU using a web-based randomisation system managed by King’s Clinical Trials Unit (KCTU). Randomisation was stratified by NHS Trust using variable block sizes. Outcome assessors were blinded to group allocation, while participants and clinicians were aware of treatment assignments. The senior trial statistician and senior trial health economist remained blinded until the final stages of analysis, and full unblinding of the junior trial statistician and trial health economist occurred only after the final database extract in June 2024.

### Interventions

The ICCS pathway is a specialised care model designed to treat young people with severe psychiatric disturbances within community settings rather than hospitals. This approach is implemented by multidisciplinary teams consisting of psychiatrists, psychologists, social workers, and nurses, offering a tailored, evidence-based treatment plan for each service user. The core of ICCS involves maintaining a low service user-to-provider ratio, typically no more than 10:1, ensuring personalised and intensive care. Teams meet regularly to coordinate and evaluate individual care plans, ensuring a collaborative and adaptive treatment approach. Interactions with service users occur mainly in community settings such as homes, schools, and cultural centres, facilitating access to natural support networks and enhancing engagement. These interactions are frequent, with multiple weekly contacts to maintain engagement and monitor progress. To provide comprehensive care, ICCS integrates psychological, pharmacological, and social interventions, including supported discharge from inpatient care, providing an alternative to inpatient admissions and outreach services. Additionally, operational standards are rigorously maintained with clearly defined criteria for admission, ongoing care, and discharge, which includes out-of-hours support and proactive involvement in hospital admissions. This model aims to provide an effective alternative to inpatient care, focusing on immediate stabilisation and long-term wellness. It supports the young person’s reintegration into their community and everyday activities, such as education and employment. The ICCS pathway was based on the characteristics defined by a consensus meeting of the investigators (13). The TAU pathway includes the standard inpatient and outpatient services available within the CAMHS framework, excluding the ICCS. TAU for those young people considered for inpatient care typically includes hospital admissions but can vary widely depending on the specific protocols and resources of the participating NHS organisations. TAU typically begins with an assessment of the young person’s mental health needs, followed by a corresponding treatment plan that could involve a combination of psychological, pharmacological, and social interventions. Inpatient care, when utilised under TAU, involves hospitalisation, aiming to stabilise the patient through intensive support and monitoring. The duration of hospital treatment varies, but it is followed by standard community treatment, which includes regular follow-up visits to outpatient services to ensure ongoing support and care continuity. Outpatient treatment under TAU may involve regular therapy sessions, medication management, and other supportive services like education about mental health, coping strategies, and relapse prevention. These services are designed to manage symptoms and support the young person’s mental health without the intensive community integration focus seen in ICCS. The control arm’s approach is a comparative standard to evaluate the effectiveness and efficiency of the more intensive, community-focused ICCS model. This comparison aims to delineate the benefits of implementing a more proactive and integrated treatment approach within the community setting instead of conventional psychiatric treatment modalities. Key intervention components are summarised in **Table 1** (TIDieR checklist).

**Table 1:** TIDieR summary of ICCS versus TAU intervention components.

| TIDieR Item | ICCS | TAU |
| --- | --- | --- |
| Why | Provide intensive, community-based crisis support to reduce time to EET and avoid admission | Deliver standard outpatient CAMHS care |
| What | Multidisciplinary home-based crisis service with daily in-person or telehealth contacts; optional day-hospital sessions | Standard CAMHS outpatient input (psychological/ pharmacological) |
| Who | Consultant psychiatrist, psychologist, community psychiatric nurse, social worker | CAMHS psychiatrist or community psychiatric nurse |
| How & where | Flexible outreach—home visits or remote contacts; patient's home or day-hospital facilities | Office-based clinic appointments at CAMHS outpatient clinics |
| When & how much | Daily contacts until clinical goals met (target within ~3 months) | Weekly or fortnightly appointments per local policy |
| Fidelity & Contamination | Fidelity tools piloted at 2 sites but not consistently collected; contamination not systematically assessed | Not monitored |

### Trial feasibility assessment

The feasibility of proceeding to a full evaluation trial was assessed at the end of the internal pilot phase. The following criterion had been pre-specified: recruitment of at least 55 participants within the first 12 months. Recruitment of ≥55 participants in 12 months was the sole progression criterion; fidelity monitoring and data completeness were tracked for internal oversight but were not prespecified as progression criteria.

### Process variables

We collected the number of contacts with mental health professionals and treatment exposure. These data were extracted from participants’ medical records. Adherence to intervention was assessed by comparing the number of treatment sessions offered with the number of treatment sessions attended.

### Primary and Secondary Outcomes

The evaluation trial’s primary outcome was the time to return to or start EET, measured from randomisation to the first day attending EET. Participants not returning to EET by the six-month follow-up were censored at their last contact date. Any return to or start of EET—regardless of duration—was counted from the date of first attendance, with engagement dates verified through clinical records and confirmation from educational or employment providers. Ambiguous cases (e.g., minimal attendance or short-term employment) were reviewed at weekly trial meetings and adjudicated on a case-by-case basis. This outcome reflects the integration of young people back into their normal activities, which is a critical indicator of recovery and social functioning. It was assessed through reports from clinical teams and educational or employment institutions to verify the young person’s engagement with EET, capturing the effectiveness of the intervention in facilitating a quicker return to daily life and productivity (10).

The secondary outcomes encompassed a broad range of measures designed to evaluate the impact of the interventions on young people’s clinical symptoms, functioning, satisfaction with services, and overall mental health. The outcomes include assessments of mental health status using the Child version of the Strengths and Difficulties Questionnaire (SDQ) and the Children’s Global Assessment Scale (CGAS). These instruments were administered at baseline and at 6 months post-randomisation to measure emotional and behavioral problems and overall mental functioning. Clinical severity was captured through the Clinical Global Impressions (CGI) and CGI Improvement Scales. These provide clinician-reported evaluations of the patient’s global functioning at baseline and changes post-intervention at 6 months. Patient satisfaction was gauged using the ChASE children self-report questionnaire, scheduled for 6 months after randomisation. General health and social functioning were tracked using Section A of the Health of the Nation Outcome Scales for Children and Adolescents (HoNOSCA) at the beginning and end of the study period. The Self-Harm Questionnaire assesses self-harm thoughts and behaviours, with evaluations conducted at the study’s start and conclusion. The length of hospital stay was quantified by the number of nights spent in psychiatric inpatient services between hospital admission and discharge, recorded from electronic patient records over the 6-month follow-up.

### Data collection and retention

To maximise follow-up, we employed flexible contact strategies—including telephone interviews, text reminders, and home or clinic visits—and enlisted treating clinicians to facilitate participant engagement with assessments.

### Health Economic Measures

Service use was measured in interviews using a modified Child and Adolescent Service Use Schedule (CA-SUS; (14)). A brief version of the CA-SUS was used at baseline, covering key service use over the previous three months as it was hypothesized that it would be difficult for participants to complete a detailed measure upon admission to a hospital experiencing a mental health crisis. Key services were high cost and/or high usage (hospital inpatient, outpatient, A&E and ambulance services, GPs, practice nurses, CAMHS workers, and therapists providing talking therapy). A detailed CA-SUS was used at 6-month follow-up covering all health and social care service use since baseline, excluding CAMHS contacts and psychiatric inpatient and day patient use, which were collected directly from medical records to maximise the accuracy of intervention data and minimise unblinding research assessors to group allocation.

The economic outcome measure was quality adjusted life years (QALYs) derived from the Child Health Utility (CHU9D) measure of health-related quality of life at baseline and 6-months post-randomisation. The CHU9D is a paediatric generic preference-based measure of health-related quality of life, consisting of nine dimensions (sad, worried, pain, annoyed, tired, homework or schoolwork, daily routine, activities, and sleep) rated using five levels (15). It is a valid and responsive utility measure for use in young people (16) (17).

### Adverse Events (AE)

Clinical teams monitored safety throughout the study. Any unfavourable or unintended signs, symptoms, or illnesses (AE) were recorded, including exacerbations of pre-existing illnesses, increased frequency or intensity of pre-existing episodic events or conditions, conditions detected after randomisation, and continuous persistent disease or symptoms present at baseline that worsen following randomisation. AEs were reported from the signing of the study consent form to the last follow-up assessment 6 months after randomisation.

### Sample size for the evaluation trial

The target sample size if the study proceeded to a full evaluation trial was initially 252 young people (126 per group). A 20% reduction in the proportion of young people not in EET (NEET) was chosen as the minimum clinically significant difference (TAU: 49% NEET, ICCS: 29% NEET). Assuming 90% power and 5% significance using a two-tailed log rank test required a sample size of 240 young people, adjusting for 5% loss to follow-up required a final sample size of 252 young people (126 per arm).

### Data Analysis

After the study stopped recruiting at the end of the pilot phase (see Results), the statistical analysis plan was adapted to estimate ICCS effect sizes based on the pilot sample. Thus, the objective of the statistical analyses became the provision of effect size estimates that can inform future evaluation studies. The objective is not to formally evaluate the benefit of ICCS. The latter is not possible due to a lack of power, and we deliberately do not report any p-values. All analyses were conducted using the intention-to-treat (ITT) principle. The primary analysis compared time to EET between ICCS and TAU using Cox proportional hazards models, adjusting for NHS Trust (SLaM, Berkshire, or Other). Kaplan-Meier survival curves were generated, and proportional hazards assumptions were assessed using Schoenfeld residuals. Secondary outcomes were analysed using linear or logistic regression models, adjusted for NHS Trust and for baseline values where appropriate. No imputation was performed for missing follow-up outcomes, and missing baseline data were handled using the missing indicator method for continuous variables. The analysis population included all randomised participants who provided baseline data. Bootstrapped confidence intervals were generated for continuous outcomes with skewed model residuals.

### Economic analysis

The economic evaluation was based on the NHS/Personal Social Services perspective preferred by NICE (18), including education-based health and social care services. Unit costs in Great British pounds for the financial year 2021-2022 were applied to individual-level service use data to calculate total costs per participant (supplementary **Table S1** and **S2**). All costs occurred within a 6-month timeframe, and discounting was therefore not applicable. QALYs were calculated using the recommended area under the curve approach (19) and applying appropriate utility weights (20). The low numbers recruited to the trial negated the feasibility of conducting an economic evaluation. We instead summarised and descriptively presented the following: (i) service use by group over the follow-up period, reporting the mean (SD) and percentage of the sample using each item; (ii) cost of service use by group over the follow-up period, reporting the mean (SD); (iii) CHU9D score and QALYs by group, reporting the mean (SD).

## Results

### Participant Recruitment and Trial Feasibility Assessment

Figure 1 illustrates the flow of participants throughout the study. At least 977 young people were screened for eligibility across multiple NHS Trusts, with 36 participants (3.7%) eventually randomised between **23/02/22** and 01/08/23. All 15 participants randomised to ICCS were included in the primary survival analysis. For the TAU group, 20 out of 21 participants contributed to this analysis. Among the four participants who withdrew from the TAU group, only one did not provide any data for either primary or secondary analyses. The feasibility of a full evaluation trial was assessed at the end of the pilot phase (August 2023). By that time, a total of 36 participants were recruited, short of the target of 55. The key reason for recruitment difficulty was that only one service could be made available for the young person in time, given the level of risk. This eliminated 206 potential participants. Thus, the progression criteria for the internal pilot phase were not met, and the study did not progress to the full RCT. This small size of the pilot sample limits the power of any group comparisons for ICCS effectiveness and cost effectiveness evaluation and thus, the sample was only analysed for the purpose of estimating the ICCS effect sizes to plan future evaluation studies. Among those screened, 897 young people were deemed not eligible. The most common reason (n = 433) was recorded as “Unwilling for CAMHS,” referring to young people or families who declined engagement with specialist child and adolescent mental health services—often due to prior negative experiences, perceived stigma, or a preference for informal or primary care support. Other exclusion categories included: “Safety risk” (n = 206), where clinical risk (e.g., suicidality or aggression) was deemed too high for ICCS to manage safely; “Below ICCS threshold” (n = 12), referring to young people who did not meet ICCS criteria such as a CGAS score ≥ 20; and “Unsuitable for randomisation” (n = 11), where clinicians were unable to maintain equipoise or where only one service pathway was practically available. No exclusions were documented due to unavailability of TAU during the pilot phase.

**Figure 1.**
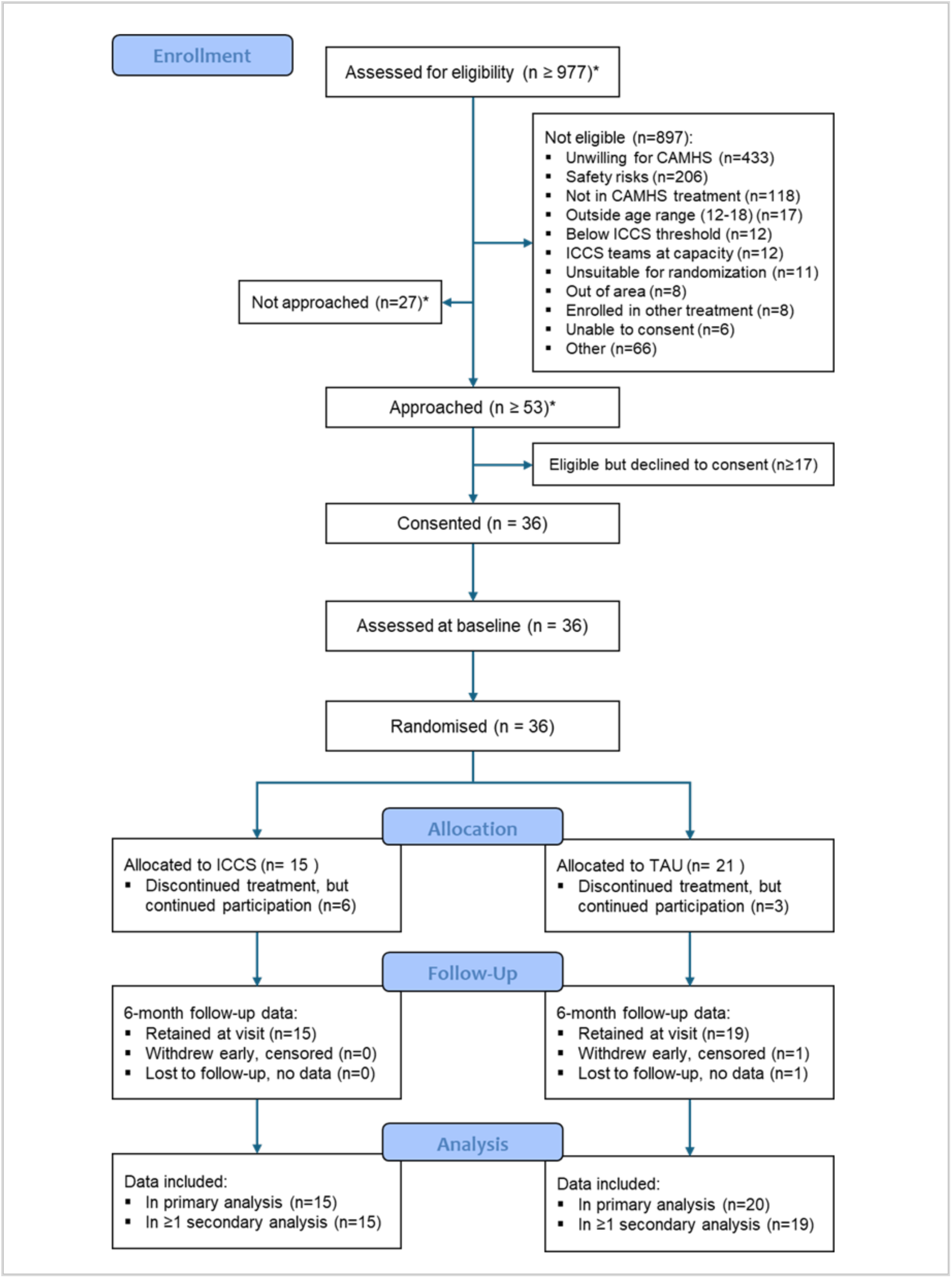
CONSORT diagram presents the participant flow in the IVY Trial, from screening (n≥977) through to randomization into ICCS (n=15) and TAU (n=21). The diagram details numbers approached, eligible, and consented, along with retention and reasons for withdrawal at the 6-month follow-up.

### Baseline characteristics of the pilot sample

The clinical and demographic characteristics of the pilot sample are summarised in **Table 2**. The majority of participants were female (77.8%), with a mean age of 15.8 years (SD = 1.3). Baseline demographic characteristics were well balanced between the two treatment arms. The ethnic composition of the sample included 47.2% White, 25% Black/African/Caribbean/Black British, and 13.9% Mixed/Multiple ethnic groups. The majority of participants (94.4%) had no prior involvement with ICCS, and none were diagnosed with borderline personality disorder at baseline. At study entry, 15 of 36 participants (41.7%) were already engaged in education, employment, or training (EET), while 21 (58.3%) were not; detailed information on the intensity of baseline engagement (e.g., full-time vs. part-time) was not collected.

**Table 2.** Baseline demographic and clinical variables by trial arm and overall.

| Baseline demographic | TAU (N=21) | ICCS (N=15) | Overall (N=36) |
| --- | --- | --- | --- |
| **Age at randomisation (n)** |  |  |  |
| mean (SD) | 15.9 (1.2) | 15.7 (1.3) | 15.8 (1.3) |
| median (IQR) | 16.4 (15.2-16.6) | 16.0 (14.6-16.9) | 16.1 (15.0-16.8) |
| **Participant sex at birth - n (%)** |  |  |  |
| Male | 5 (23.8%) | 3 (20.0%) | 8 (22.2%) |
| Female | 16 (76.2%) | 12 (80.0%) | 28 (77.8%) |
| **Ethnic group - n (%)** |  |  |  |
| White | 11 (52.4%) | 6 (40.0%) | 17 (47.2%) |
| Mixed/Multiple ethnic groups | 1 (4.8%) | 4 (26.7%) | 5 (13.9%) |
| Asian/Asian British | 3 (14.3%) | 1 (6.7%) | 4 (11.1%) |
| Black/ African/Caribbean/Black British | 6 (28.6%) | 3 (20.0%) | 9 (25.0%) |
| Other ethnic group | 0 (0.0%) | 1 (6.7%) | 1 (2.8%) |
| **IMD Decile - n (%)** |  |  |  |
| 2 - more deprived | 2 (9.5%) | 2 (13.3%) | 4 (11.1%) |
| 3 | 4 (19.0%) | 2 (13.3%) | 6 (16.7%) |
| 4 | 3 (14.3%) | 0 (0.0%) | 3 (8.3%) |
| 5 | 1 (4.8%) | 2 (13.3%) | 3 (8.3%) |
| 6 | 2 (9.5%) | 1 (6.7%) | 3 (8.3%) |
| 7 | 1 (4.8%) | 1 (6.7%) | 2 (5.6%) |
| 8 | 3 (14.3%) | 1 (6.7%) | 4 (11.1%) |
| 9 | 3 (14.3%) | 1 (6.7%) | 4 (11.1%) |
| 10 - least deprived | 2 (9.5%) | 5 (33.3%) | 7 (19.4%) |
| **Country of residence - n (%)** |  |  |  |
| England | 21 (100.0%) | 15 (100.0%) | 36 (100.0%) |
| **Is self-reported gender same as sex at birth - n (%)** |  |  |  |
| No | 2 (9.5%) | 0 (0.0%) | 2 (5.6%) |
| Yes | 19 (90.5%) | 15 (100.0%) | 34 (94.4%) |
| **Borderline Personality Disorder Diagnosis - n (%)** |  |  |  |
| No | 21 (100.0%) | 15 (100.0%) | 36 (100.0%) |
| **Psychosis Diagnosis - n (%)** |  |  |  |
| No | 17 (81.0%) | 14 (93.3%) | 31 (86.1%) |
| Yes | 4 (19.0%) | 1 (6.7%) | 5 (13.9%) |
| **Autism Spectrum Disorder Diagnosis - n (%)** |  |  |  |
| No | 18 (85.7%) | 11 (73.3%) | 29 (80.6%) |
| Yes | 3 (14.3%) | 4 (26.7%) | 7 (19.4%) |
| **Previous input from ICCS - n (%)** |  |  |  |
| No | 20 (95.2%) | 14 (93.3%) | 34 (94.4%) |
| Yes | 1 (4.8%) | 1 (6.7%) | 2 (5.6%) |

### Data completeness

Follow-up data for the primary EET outcome were available for 85% of participants, with up to 30% missingness on secondary outcomes.

### Treatment exposure and adherence

The total number of mental health contacts and treatment exposure are summarised in **Tables S3 and S4**. The median number of patient contacts with most types of mental health workers was 0, suggesting that most IVY participants did not have many mental health contacts over the observation period. There was a good adherence to ICCS treatment. Young people were offered a median of 14.0 (IQR 6.0-18.0) ICCS appointments, of which a median of 11.0 (IQR 6.0-16.0) were attended. In addition to the ICCS treatment, young people in the ICCS arm were offered a median of 5.0 (IQR 1.0-13.0) standard community treatment sessions, and they attended a median of 5.0 (IQR 1.0-8.0) of these sessions. In TAU, young people were offered a median of 8.0 (IQR 2.5-15.0) standard community treatment sessions, of which a median of 5.5 (IQR 2.5-9.5) were attended. Despite TAU permitting inpatient care, very few TAU participants experienced any hospital admission during follow-up

### ICCS effect sizes in terms of trial outcomes

The primary trial outcome was the time to start or return to EET. Among the 36 participants, 30 (83.3%) started or returned to EET within the 6-month follow-up period. The median unadjusted time to EET was 9 days overall (interquartile range [IQR]: 1-49). Participants in the ICCS arm had a shorter median unadjusted time to EET (6 days, IQR: 1-35) than those in the TAU arm (12 days, IQR: 2-84), though the interquartile ranges overlapped. The unadjusted Kaplan-Meier survival analysis (Figure 2) showed a faster transition to EET in the ICCS arm compared to TAU. After adjusting for NHS Trust, we estimate that allocation to ICCS is associated with a higher probability of returning to /starting EET in our sample (HR: 1.34, 95% CI: 0.63 - 2.86).

**Figure 2.**
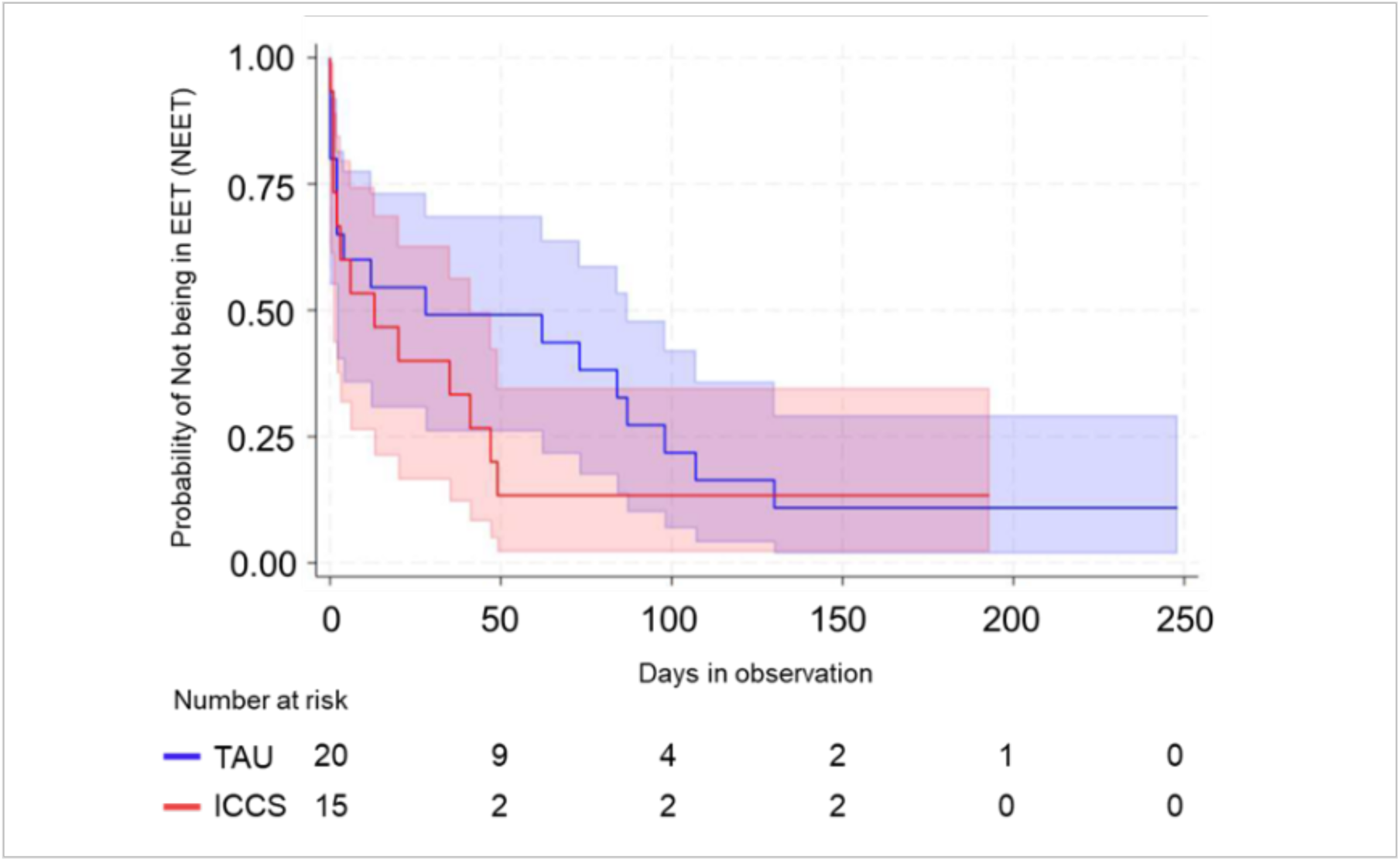
Kaplan-Meier curves depict survival probabilities, where “surviving” is defined as not engaging in employment, education, or training (NEET) for participants in the ICCS and TAU arms. All data are included following the intention-to-treat principle. The plot accounts for five participants who started/returned to EET on the same day they were randomized, shown as minimal event times of 0.001 days.

Effect size estimates for the ten secondary trial outcomes are shown in **Table 3**. This shows that all secondary outcomes in our pilot sample improved under ICCS. In the pilot sample, allocation to ICCS was associated with lower/improved SDQ scores, higher/improved CGAS ratings, lower/improved HoNOSCA Section A scores and higher/better service satisfaction scores. No clear improvements were seen between ICCS and TAU in the odds of reporting 5+ episodes of self-harm or lengthening time in EET. Nights in hospital were omitted from the analysis because there were only 4 participants who reported being admitted to a psychiatric inpatient ward. In the pilot sample, participants randomised to ICCS had lower odds of having a 1-point increase/worsening in CGI Illness Severity rating. However, it should be noted again that we were not powered to formally assess the existence of any group differences.

**Table 3.**
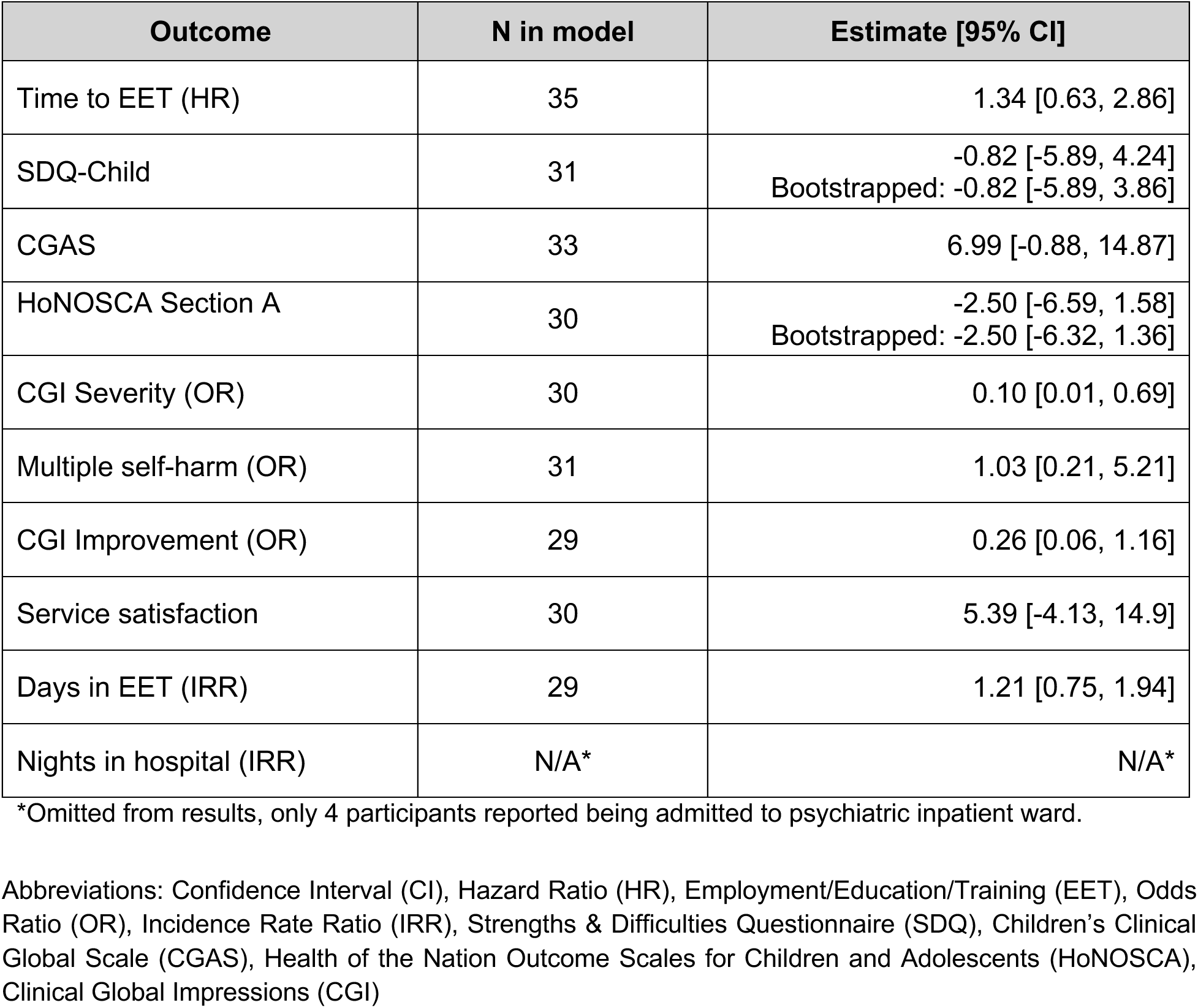
The effect of treatment (estimated difference between ICCS and TAU) on the primary and secondary outcomes, using TAU as the reference group and adjusting for NHS Trust and baseline values where appropriate.

### Safety and Adverse Events

A total of 12 safety events, 5 in ICCS and 7 in TAU were reported during the study period, including three serious adverse events (SAEs) in the TAU and three in ICCS. All six SAEs were of mild or moderate severity, and only one SAE was reported as probably related to the study intervention.

### Health economics

Full economic data (full-service use and CHU9D data at baseline and 6-month follow-up) were available for 28 participants (78% of all recruited participants). An equal number of participants with full economic data was available in each group (n=14 ICCS; n=14 TAU). Extensive utilization of hospital services for mental health reasons was observed across both study groups before the trial’s commencement, as shown in supplementary **Table S5**. Notably, 79% of participants in both groups utilised inpatient admissions and emergency services, while outpatient appointments were accessed by 32%. Engagement with CAMHS workers was also substantial, reported by 71% of all participants, indicating a critical need for mental health support in this population.

Follow-up data are reported in **Tables S6 and S7** (use of intervention services and use of all other health and social care services, respectively). In terms of intervention use (**Table S6**), as would be expected, the ICCS group used more ICCS-specific interventions, whilst the control group used more of many, although not all, of the non-ICCS-specific services. Direct comparison of individual services is not meaningful, given randomisation to ICCS or TAU, but in aggregate, contacts excluding inpatient admissions were higher in the ICCS group (mean 24.21, SD 18.99) compared to TAU (mean 9.57, SD 10.06), whilst psychiatric inpatient nights were lower in the ICCS group (mean 3.21, SD 12.03) compared to TAU (mean 15.07, SD 38.42). The use of all services was low with no obvious patterns of differences between groups.

The costs of all services used between baseline and follow-up are reported in **Table 4**. During the 6-month follow-up period, the total costs related to intervention/control service use were lower for the ICCS group (mean £5,640, SD 10,074), as compared to TAU (mean £13,526, SD 31,702)), due primarily to lower costs incurred from psychiatric inpatient nights (mean £2,515 ICCS versus mean £11,794 TAU). Follow-up costs from the CA-SUS were also lower for ICCS (mean £1,423, SD 2,553) compared to TAU (mean £1,629, SD 2,191), resulting in lower total costs for the ICCS group (mean £7,063, SD 10,605) compared with the TAU group (£15,155, SD 31,560). The calculation of QALYs based on the CHU9D utility scores from baseline to 6-month follow-up revealed no significant differences in health-related quality of life between the groups (**Table 5**).

**Table 4.** Mean Costs (£) Per Participant Over the 6 Month Follow-up.

|  | All (n=28) |  | ICCS (n=14) |  | TAU (n=14) |  |
| --- | --- | --- | --- | --- | --- | --- |
| Service | Mean £ (SD) | Range £ | Mean £ (SD) | Range £ | Mean £ (SD) | Range £ |
| **Hospital** |  |  |  |  |  |  |
| PH Inpatient | 95 (369) | 0-1,771 | 190 (513) | 0-1,771 | 0 (0) | n/a |
| PH Day Case | 110 (584) | 0-3,088 | 221 (825) | 0-3,088 | 0 (0) | n/a |
| PH Outpatient | 292 (756) | 0-3,160 | 301 (838) | 0-3,160 | 282 (696) | 0-2,633 |
| A&E Attendance | 77 (132) | 0-432 | 82 (135) | 0-432 | 72 (135) | 0-432 |
| Ambulance Services | 25 (131) | 0-695 | 0 (0) | n/a | 50 (186) | 0-695 |
| Health-based Place of safety | 44 (234) | 0-1,236 | 0 (0) | n/a | 88 (330) | 0-1,236 |
| Total Hospital | 643 (1,742) | 0-8,451 | 794 (2,227) | 0-8,451 | 492 (1,139) | 0-4,157 |
| **Community** |  |  |  |  |  |  |
| GP | 71 (80) | 0-369 | 59 (66) | 0-231 | 82 (93) | 0-369 |
| Practice nurse | 3 (7) | 0-23 | 3 (5) | 0-12 | 4 (9) | 0-23 |
| Other nurse | 21 (80) | 0-397 | 12 (44) | 0-165 | 30 (106) | 0-397 |
| Therapist Talking therapy | 583 (944) | 0-3,960 | 377 (615) | 0-1,650 | 790 (1,176) | 0-3,960 |
| Education MH Practitioner | 85 (225) | 0-986 | 88 (189) | 0-657 | 82 (263) | 0-986 |
| Community Paediatrician | 50 (184) | 0-700 | 50 (187) | 0-700 | 50 (187) | 0-700 |
| SEN Coordinator | 5 (20) | 0-75 | 5 (20) | 0-75 | 5 (20) | 0-75 |
| Social worker | 44 (126) | 0-614 | 30 (78) | 0-236 | 57 (163) | 0-614 |
| OT | 12 (46) | 0-210 | 0 (0) | n/a | 24 (63) | 0-210 |
| SLT | 2 (8) | 0-42 | 3 (11) | 0-42 | 0 (0) | n/a |
| Drug/alcohol Support worker | 6 (30) | 0-160 | 0 (0) | n/a | 11 (43) | 0-160 |
| Helpline/advice Service | 1 (3) | 0-12 | 2 (4) | 0-12 | 1 (2) | 0-8 |
| Total Community | 883 (1,001) | 0-4,727 | 629 (660) | 0-1,742 | 1,137 (1,227) | 104-4,727 |
| **Total** |  |  |  |  |  |  |
| 6-month CA-SUS cost | 1,526 (2,337) | 140-10,061 | 1,423 (2,553) | 144-10,061 | 1,629 (2,191) | 141-8,884 |
| ICCS/TAU Cost | 9,583 (23,428) | 0-96,391 | 5,640 (10,074) | 217-36,432 | 13,526 (31,702) | 0-96,391 |
| **6-month Follow-up cost** | 11,109 (23,467) | 248-98,277 | 7,063 (10,605) | 454-38,318 | 15,155 (31,560) | 248-98,277 |

MH Mental Health; PH Physical Health; A&E Accident and Emergency; GP General Practitioner; SEN Special Education Needs; OT Occupational Therapist; SLT Speech and Language Therapist; CA-SUS Child and Adolescent Service Use Schedule.

**Table 5.** Mean utility scores and 6-month QALYs per participant.

|  | All (n=28) |  | ICCS (n=14) |  | TAU (n=14) |  |
| --- | --- | --- | --- | --- | --- | --- |
|  | Mean (SD) | Range | Mean (SD) | Range | Mean (SD) | Range |
| CHU9D |  |  |  |  |  |  |
| Baseline utility | 0.718<br>(0.151) | 0.415-1.000 | 0.709<br>(0.141) | 0.415-0.963 | 0.727<br>(0.166) | 0.532-1.000 |
| 6-month utility | 0.740<br>(0.127) | 0.445-1.000 | 0.740<br>(0.118) | 0.445-0.926 | 0.740<br>(0.139) | 0.507-1.000 |
| 6-month QALYs | 0.364<br>(0.058) | 0.267-0.488 | 0.362<br>(0.055) | 0.267-0.456 | 0.367<br>(0.064) | 0.290-0.488 |

CHU9D Child Health Utility-9 Dimensions; QALY Quality-Adjusted Life Year.

## Discussion

### Main Findings

This study represents the first attempt at a trial conducted in the UK to evaluate the effectiveness of ICCS compared to TAU for young people with severe psychiatric disorders considered for inpatient admissions. The study did not achieve the progression criteria of the internal pilot phase, and it was concluded that conducting an evaluation trial in the UK was not feasible at this stage, the main difficulty being capacity issues that prevented services from offering two alternative treatment pathways at the same time, an issue that was made worse by increased demand for mental health support for young people after the COVID period.

To aid future evaluation studies, we estimate the effect of ICCS compared to TAU in terms of planned trial outcomes. While underpowered for any formal evaluation, the results from the pilot sample are promising and support further investigation of ICCS benefits. Participants in the ICCS arm demonstrated a quicker return to EET, with a median unadjusted time to EET of 6 days, compared to 12 days in the TAU arm. This may indicate that ICCS effectively facilitates the reintegration of young people into educational and work environments following psychiatric emergencies. The limited data highlight the need for a larger, more robust study to formally assess the effectiveness of ICCS and to explore the mechanisms through which ICCS may expedite reintegration into societal roles for young people following psychiatric crises. There was no evidence of an increase in adverse events with ICCS.

The development of the current ICCS model was significantly influenced by earlier findings from our research group (21), which compared the effectiveness of Supported Discharge Service, which represents one of three key aspects of ICCS, with standard inpatient care. Despite showing no significant differences in functional impairment and clinical outcomes, there were significant differences in educational outcomes and self-harm favouring ICCS. These findings underpinned the rationale for developing and refining community-based care models. Previous evidence of the effectiveness of one component of ICCS, together with effect size estimates in this study, might indicate that ICCS has more benefits than TAU. Given the pilot’s limited sample size, we do not interpret observed treatment effect sizes and instead focus on key feasibility metrics— recruitment barriers, data completeness (15% missing primary outcome; 30% secondary), and fidelity monitoring challenges—which are essential for planning a definitive trial. To address these feasibility constraints, future evaluations could employ alternative designs such as stepped-wedge cluster trials to improve recruitment flexibility, cluster-randomised designs to reduce contamination, or hybrid implementation– effectiveness trials to assess both ICCS delivery and clinical outcomes concurrently (22, 23).

While ICCS is already implemented across the NHS, the substantial ineligibility rate of 91.8% observed in our study underscores critical areas for potentially optimising these services. Whereas most ineligible young people clearly needed one of the existing services, other factors played an important role. They included reluctance to engage with CAMHS, complex risk profiles, and logistical constraints like local ICCS team capacities and geographical limitations, pointing to potential barriers that may restrict access to care for adolescents with psychiatric emergencies. Acknowledging these barriers not only highlights the need for continuous improvement in the delivery of community-based care but also calls for a deeper investigation into how these services can be made more inclusive and responsive to the needs of all patients.

In this small pilot sample, the ICCS group demonstrated similar QALYs to the TAU group alongside lower overall health and social care costs over the study period, which was largely attributable to lower utilisation of high-cost psychiatric inpatient services. Despite this cost difference in favour of ICCS, inferences about costs and cost-effectiveness could not be made and no adjustments were made for baseline differences, so the presented data should be treated cautiously and used only to generate hypotheses for future research.

### ICCS Implementation in this study

The implementation of ICCS in our study was conservative. We required all participating ICCS to have access to a day hospital service, which excluded a significant number of services. This conservative approach ensured a consistent standard of care across all ICCS settings in the study. It may have limited the generalisability of our findings to other

ICCS models that do not incorporate day hospital care. This decision was made to strengthen the comparability of ICCS to more structured services like inpatient care, but it restricted the diversity of ICCS approaches that could be explored in this trial.

### Adherence to Treatment

Unlike many other studies of psychiatric treatments, where adherence is often suboptimal, we observed good adherence to both ICCS and TAU. This was particularly notable given the severe psychiatric disorders in our sample. The active engagement of young people and their families in treatment may reflect the high level of need and the tailored, intensive nature of ICCS. However, it should also be noted that adherence may have been facilitated by the strong relationships between clinical teams and participants, which could be less pronounced in larger, more diverse trials.

### Real-Life Study Design

Our study was conducted in real-life clinical services, which adds to the ecological validity of the findings. However, this also presented several challenges, notably the impact of the COVID-19 pandemic, which affected service delivery and recruitment. The pandemic led to increased demand for mental health services, which may have influenced the capacity of clinical teams to refer young people to the trial and impacted their decision-making regarding the most appropriate care pathway for each individual. Despite these challenges, our study maintained a high follow-up rate for the primary outcome measure, which strengthened the reliability of the data we were able to collect.

Recruitment was one of the main challenges of this trial, with several obstacles uncovered during the feasibility phase. One significant barrier was the determination of clinical teams about which pathway (ICCS or TAU) was most suitable for individual young people, given the risk profile. In many cases, only one pathway was available and teams based their decisions on limited evidence and clinical experience rather than randomisation, which led to slower recruitment rates. Additionally, the high level of complexity and acuity of the participants’ conditions may have influenced clinicians’ willingness to randomise them into different care pathways, reflecting real-world concerns about service suitability.

### Limitations

The most significant limitation of this pilot study is the very small sample size, which was a direct result of recruitment difficulties. As a result, we were unable to draw any inferences about the effectiveness or cost-effectiveness of ICCS relative to TAU. Additionally, it was not possible to blind participants to the intervention they were receiving, which may have introduced bias in self-reported outcomes such as satisfaction and functioning. We also acknowledge that specific reasons for referral to ICCS or inpatient admission were not systematically recorded, limiting our ability to characterise the full clinical context for eligibility decisions. Future trials should incorporate structured collection of referral indications across sites. While fidelity monitoring tools were piloted at two sites, we did not achieve consistent fidelity data collection across all sites nor formally assess potential contamination between study arms; future work should incorporate these measures to bolster internal validity. The low number of TAU hospitalisations—which may reflect bed shortages, clinician reluctance to admit, or early symptom resolution—limits our ability to assess ICCS’s hospital-avoidance function in this pilot. We also note that 41.7% of participants were already engaged in education, employment, or training at baseline; without data on the intensity of that engagement (e.g., full-vs. part-time), our time-to-EET outcome may be influenced by pre-existing participation, and future studies should record both the presence and extent of baseline EET involvement. We observed 15% missing data for the primary EET outcome and up to 30% missingness on secondary measures. Although mitigated by flexible contact and clinician support, future trials should employ digital data-capture platforms and automated reminders to further improve retention and minimise missing data.

### Strengths

Despite the limitations, this study has several strengths. It is the first randomised study in the UK to directly compare ICCS with existing services for young people with severe psychiatric disorders, providing important preliminary data in an area with limited research. Our sample was diverse, both in terms of demographics and psychiatric diagnoses, and the real-world clinical setting enhances the generalizability of the findings to everyday practice. Moreover, the study demonstrated high adherence rates and good participant engagement, which is promising for future trials involving this population.

### Implications

In conclusion, this pilot RCT suggests that ICCS may be a promising intervention for young people with severe psychiatric disorders. Although the sample size was insufficient to draw definitive conclusions, the effect size estimates are promising and previous studies indicate that one aspect of ICCS, Supported Discharge Service, appears to be efficacious in terms of school reintegration and reducing self-harm. Future studies with larger sample sizes are necessary to confirm the findings of this study and explore the full potential of ICCS as an alternative to inpatient and other community-based services for young people with severe mental health needs. An ongoing post-implementation evaluation should be done for those areas that implement ICCS possibly utilising routinely collected outcome data. This study also provides important information to support such future research, including insights into recruitment challenges and effect size estimates that will inform the design of future evaluation studies.

## Supplementary Materials

Supplementary material is available

## Declaration

### Ethics approval and consent to participate

Ethical approval for this study was granted by the West Midlands and Black Country Research Ethics Committee (REC reference: 20/WM/0069). The trial was prospectively registered with the ISRCTN registry (ISRCTN42999542; registration date: 29 April 2020). All participants, or their legal guardians for those under 16 years of age, provided written informed consent prior to enrolment. The study was conducted in accordance with the Declaration of Helsinki (1975), as revised in 2013, and complied with all relevant national and institutional ethical standards.

### Consent for publication

Not applicable

### Availability of data and materials

All data generated or analysed during this study are included in this published article and its supplementary materials

## Authors’ contributions

DO, SB, TZ, and SL conceived the study and obtained funding. SL and PC conducted the statistical analyses. SB, MH, and ET performed the health economic evaluation. All authors contributed to the interpretation of findings, critically reviewed the manuscript for intellectual content, and approved the final version.

## Data Availability

All data produced in the present study are available upon reasonable request to the author

## Acknowledgements

The authors would like to thank the young people and families who participated in the study during an exceptionally challenging period in their lives. We are also grateful to the NHS Trusts and Health Boards that supported recruitment and delivery of the study.

